# Assessing Emergency Clinicians’ Needs from Trauma Clinical Guidance: A Mixed-Methods Study

**DOI:** 10.64898/2026.02.16.26346423

**Authors:** Lindsay Fountain, Kayla Corredera-Wells, Nicholas Cozzi, Jeffrey M. Goodloe, Jenny M. Guido, Alyssa B. Johnson, Christopher S. Kang, Thomas McNally, Andrea L. Nevedal, James E. Winslow, Gabriela Zavala Wong, Lacey N. LaGrone

## Abstract

**Background:** In the United States, emergency clinicians are often the first to care for injured patients in the hospital setting. To better understand end-user needs, we evaluated emergency clinician priorities and preferences in accessing, interpreting, and applying trauma clinical guidance.

**Methods:** Emergency clinicians were recruited via email for semi-structured video conference interviews. Rapid directed qualitative analysis of interview notes and audio recordings yielded initial insights about guidance barriers and facilitators. A subsequent quantitative survey was developed and distributed via email to members of relevant professional associations. Survey results were analyzed using descriptive and inferential statistics.

**Results:** Twelve emergency clinicians participated in interviews. 154 eligible participants responded to the survey. Clinicians expressed support for trauma clinical guidance overall but often find resources lacking. Barriers to guidance usage include lack of awareness, difficulty locating guidance, and cumbersome design. Clinical guidance should be objective, concise, updated, and easy-to-use at bedside. The strongest determinant of guidance usability was being quickly understood in a time-pressured situation. Clinicians prefer to access guidance through mobile applications or multi-modal channels. Rural clinicians reported additional difficulties in staffing and having resources needed to follow guidance.

**Conclusion:** When developing trauma clinical guidance, the trauma community should continue to consider the variety of end users and clinical settings, including emergency clinicians.

Developing mobile device-friendly, quickly understandable guidance should be a priority for authors of trauma clinical guidance.

## Introduction

Studies show that contemporary clinical guidance improves patient outcomes, improving care quality as it lowers morbidity, mortality, and patient harm.^1, 2, 3^ However, clinical guidance today is often fragmented, heterogenous, and inconsistently applied in emergency departments (EDs) in the United States, with guidance adherence ranging from 40-60%.^4, 5^ Emergency clinicians face multifaceted barriers to using clinical guidance, including that concerning trauma care. Emergency clinicians are often amongst the first to care for injured patients, particularly outside of Level I and II trauma centers. Urgency exists around the need to improve trauma care, with US trauma mortality rates rising 91% from 2000 to 2020.^6^ This growing problem creates opportunities to reduce trauma fatalities overall and for emergency clinicians to utilize trauma clinical guidance towards achieving better outcomes.

Deeper understanding of emergency clinicians’ needs from trauma clinical guidance can enable those clinicians to more consistently utilize and find satisfaction with guidance. A previous study identified clarity of presentation and rigor of development as the guidance characteristics with the highest correlation with emergency clinicians’ overall satisfaction with clinical guidance.^7^ Other recent research at an academic-affiliated, urban Level I trauma center identified key barriers to guidance usage in the ED and developed an electronic database of guidance with visually-appealing templates, increasing usage of and satisfaction with guidance in that ED.^8^ However, further research is needed into the preferences of emergency clinicians working in different practice settings, as well as preferences specific to guidance concerning care for injured patients. This exploratory, sequential mixed-methods study aims to determine emergency clinicians’ needs and preferences for trauma clinical guidance.

## Methods

### Study Design and Setting

This study employed an exploratory, sequential mixed-methods design, using semi-structured interviews followed by a national, electronic survey of US-based emergency clinicians. Video conference interviews took place in August and September 2024. Insights gained from those interviews informed the development of an online survey designed to further explore and validate the results identified in interviews. The survey was disseminated during February and March 2025 via email through emergency medicine (EM) professional associations. See Supplemental Items 1 and 2 for additional methods.

This study was reviewed by the Colorado Multiple Institutional Review Board, CB F490; COMIRB #: 24-0047 and determined exempt. Interviewees provided verbal consent. Respondents completed a consent form at the start of the survey. No compensation was given for participation. Only those doing direct interview analysis (LF, KCW) knew the identifying information of the interview participants. That information was abstracted before sharing data or analysis further within the team. Survey participants’ anonymity was protected by collecting limited identifiable information, analyzing responses at the characteristic rather than individual level, and using existing protections within the survey data capture tool.

### Selection of Participants

Eligible participants were defined as emergency clinicians, including physicians and advanced practice providers, actively practicing in the US at the time of interview or survey. Purposeful criterion and snowball sampling were used to recruit interview participants.^9^ To diversify the sample, we aimed to include emergency clinicians across gender, race, ethnicity, age, geographic region (CDC definition), trauma center level, clinician profession, years since completing training, and level of urbanity.^10^ Clinicians were recruited for interviews via email outreach, leveraging the research team’s network of professional contacts and, as the research progressed, referrals from past participants. Although some participants were known to the project team at the time, none had preexisting relationships with the interviewer, and their identities were blinded to the project team as analysis began.

Thirty-four individuals were contacted, twenty-two declined, and twelve completed interviews. The concept of information power, which indicates that “the more information the sample holds, relevant for the actual study, the lower number of participants is needed,” was chosen to inform sample size needs for interviews.^11^ Although saturation is commonly discussed in qualitative methods, it was not appropriate in this study that did not aim to generate theory.^12^ The criterion sample size of twelve was sufficient and yielded higher information power given the specific aims (feedback on specific trauma resources), dense sample specificity (practicing emergency clinicians with expert knowledge), and strong dialogue with participants (expertise in EM topic, qualitative methods, and focused discussions).^13^

Survey participant recruitment utilized EM professional associations to disseminate the online survey via email. Voluntary response sampling guided survey data collection. The Society of Emergency Medicine PAs shared the survey with its entire membership, and the American College of Emergency Physicians shared the survey with members of its Rural, Trauma and Injury Prevention, and Medical Directors sections.

### Measurements and Data Analysis

The interview guide included open-ended questions about clinician preferences for guidance (Supplemental Item 3). Clinicians also commented on two examples of existing trauma clinical guidance, which features protocols routinely used in the care of injured patients and provided a variety of visual styles for participants to react to. The protocols shared were the 2023

American Association for the Surgery of Trauma (AAST)/American College of Surgeons Committee on Trauma (ACS-CoT) Protocol for Damage Control Resuscitation of the Trauma Patient (Figure 1) and the University of California San Francisco (UCSF) Adult Massive Transfusion Protocol (Figure 2).^14,15^ Indicators of authorship removed before sharing with participants. Example protocols were emailed to participants at least 48 hours prior to their interview for review.

**Figure 1.**
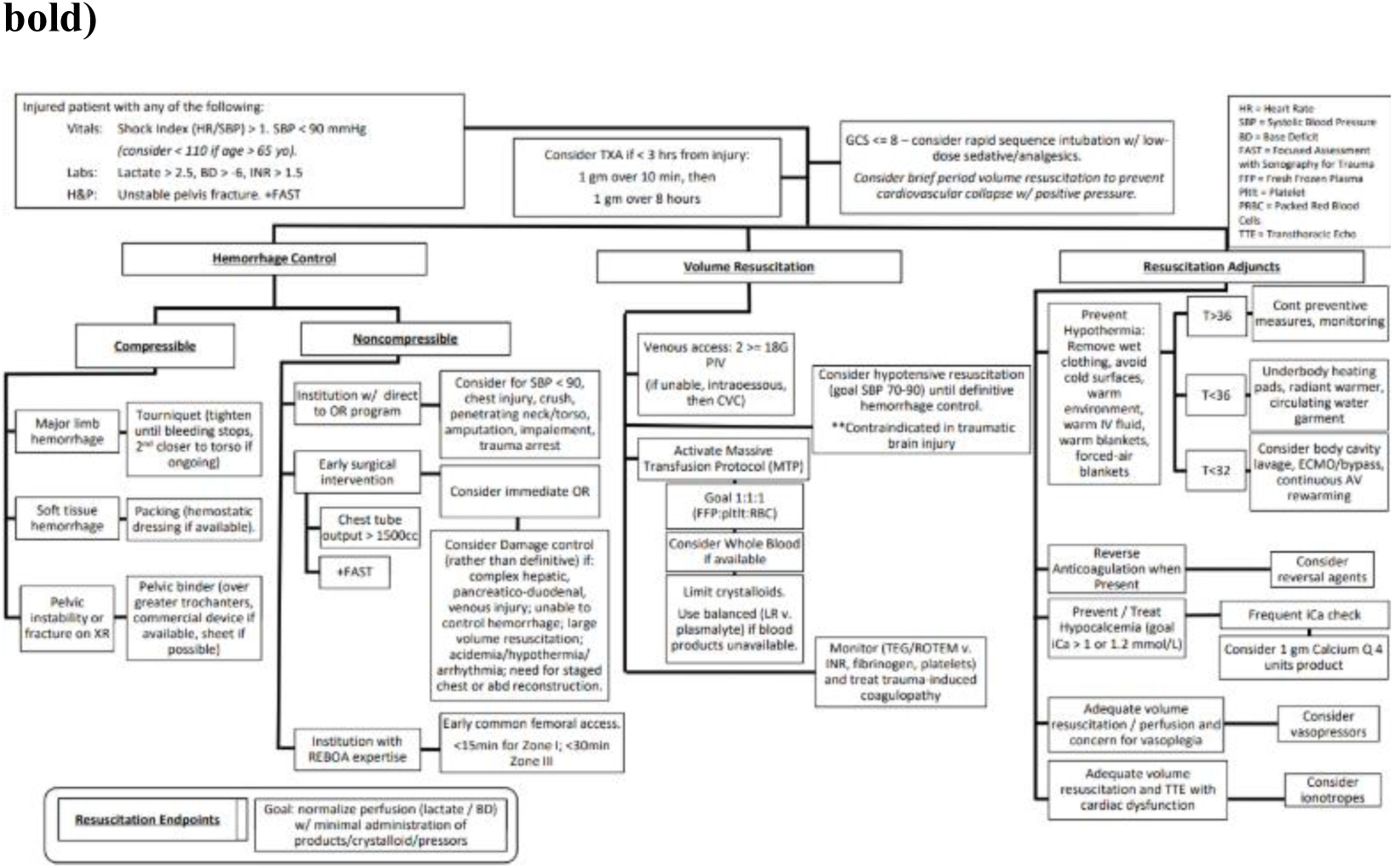
Version of AAST/ACS-CoT Protocol for Damage Control Resuscitation of the Trauma Patient shared with participants.

**Figure 2.**
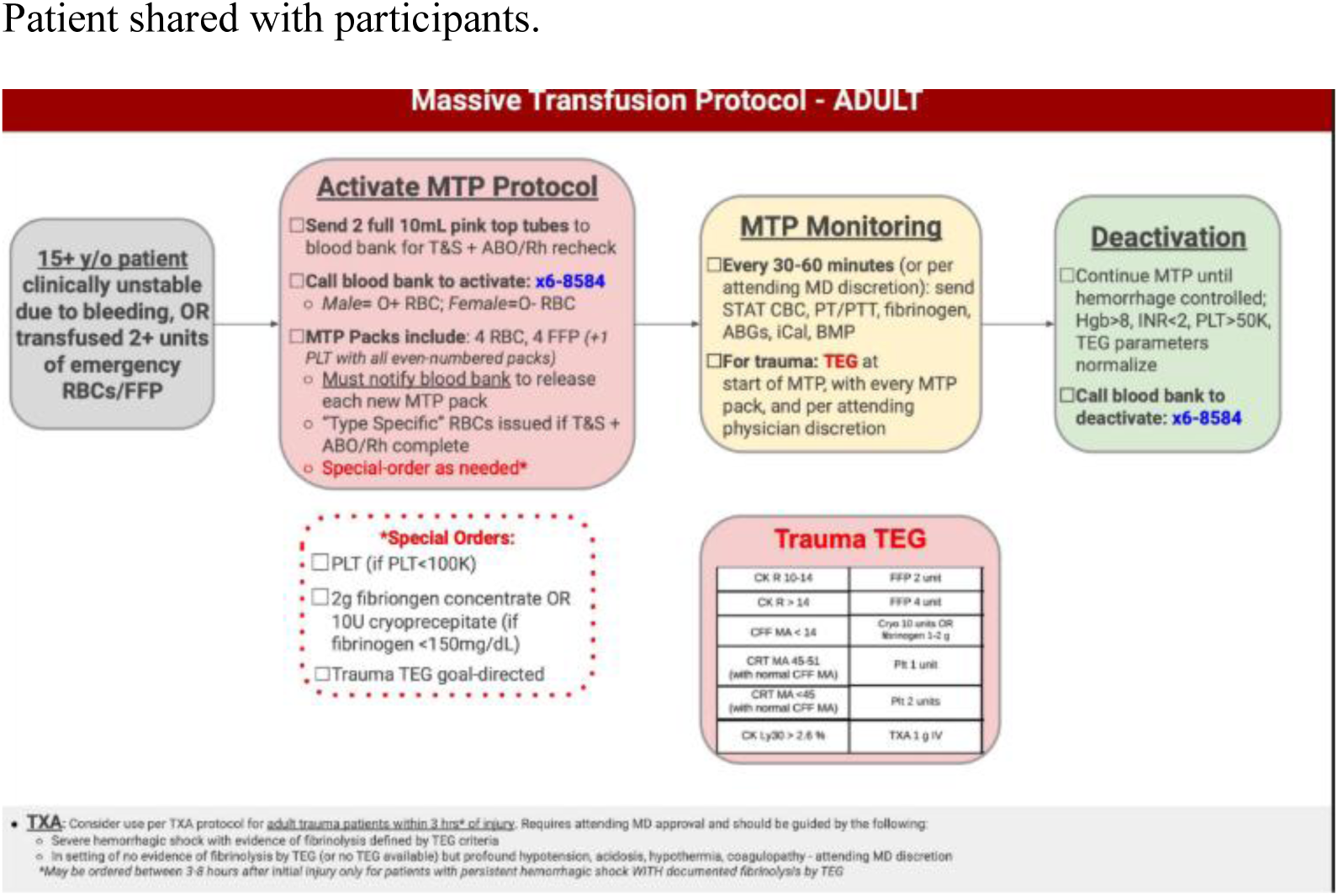
Version of UCSF Adult Massive Transfusion Protocol shared with participants.

Interviews were audio recorded and reviewed by two analysts (LF, KCW) using rapid directed qualitative analysis, with methodologic expertise provided by a medical anthropologist (AN) with significant experience in rapid qualitative methods.^16,17,18^ The interviewer and primary analyst (LF) took notes during interviews using a template created in Microsoft Office Word.

The interviewer asked clarifying questions as needed during interviews, and participants were encouraged to provide candid feedback. The primary analyst then used these notes to draft an interview summary immediately after each interview. The secondary analyst (KCW) listened to the audio recordings of each interview to verify the accuracy of and edit each interview summary. The primary analyst then organized the summarized data and exemplar quotes into a Microsoft Office Excel matrix containing rows for each participant and columns for domains for each interview question. A Microsoft Word summary was created by the primary analyst for each interview domain, enabling comparison of participant responses.

The cross-sectional online survey was designed to further explore the insights identified during interviews. Survey data were collected and managed using REDCap electronic data capture tools hosted at the University of Colorado.^19^ Survey data was analyzed using Microsoft Excel and STATANow BE version 18.5. Descriptive statistics as well as regression were used to analyze survey data. Multivariable linear regression was used to determine if participant characteristics (Table 2) predicted key continuous outcomes (e.g., frequency of guidance usage). Binary logistic regression was used to determine if the same participant characteristics predicted noncontinuous outcomes of interest (e.g., identifying cost as a top two barrier preventing guidance usage). For all regression analyses, independent variables were transformed into indicator variables to maintain sufficient sample sizes while still isolating groups of interest and their impact on key outcomes (Table 3). For example, responses from those working in level I trauma centers were compared to those working at all other trauma center levels combined.

**Table 1.** Interview participants self-reported characteristics.

| **Characteristic** | **Item** | **Count** | **Percentage (%)** |
| --- | --- | --- | --- |
| Clinician profession | Physician | 10 | 83 |
|  | PA | 2 | 17 |
| Clinical duties | Bedside | 9 | 75 |
|  | Administrator | 3 | 25 |
| Years of experience | <10 years | 4 | 33 |
|  | 10-20 years | 3 | 25 |
|  | 20+ years | 5 | 42 |
| Trauma center level | Level I | 4 | 33 |
|  | Level II | 3 | 25 |
|  | Level III | 4 | 33 |
|  | Level IV | 2 | 16 |
|  | Level V | 1 | 8 |
| Degree of urbanity | Urban | 6 | 50 |
|  | Suburban | 4 | 33 |
|  | Rural | 3 | 25 |
|  | Frontier | 1 | 8 |
| Geographic region | Northeast | 2 | 16 |
|  | South | 2 | 16 |
|  | Midwest | 1 | 8 |
|  | West | 7 | 50 |
| Gender | Female | 6 | 50 |
|  | Male | 6 | 50 |
| Race and/or ethnicity | Asian | 1 | 8 |
|  | Black | 3 | 25 |
|  | Hispanic | 2 | 16 |
|  | White | 6 | 50 |

**Table 2.** Survey respondent self-reported characteristics.

| **Characteristic** | **Item** | **Count** | **Percentage (%)** |
| --- | --- | --- | --- |
| Clinician profession<br>(n = 132) | Attending | 48 | 36 |
|  | Nurse practitioner | 3 | 2 |
|  | PA | 81 | 61 |
| Years of experience<br>(n = 144) | <10 years | 44 | 31 |
|  | 10-20 years | 41 | 28 |
|  | 20+ years | 59 | 41 |
| Trauma center level<br>(n = 154) | Level I | 40 | 26 |
|  | Level II | 29 | 19 |
|  | Level III | 23 | 15 |
|  | Level IV | 21 | 14 |
|  | Level V | 9 | 6 |
|  | Non-Level I center, unspecified level | 31 | 20 |
| Degree of urbanity<br>(n = 154) | Urban | 51 | 33 |
|  | Suburban | 50 | 32 |
|  | Rural | 63 | 41 |
|  | Frontier | 4 | 3 |
|  | Did not specify | 1 | 1 |
| Gender<br>(n = 143) | Female | 61 | 43 |
|  | Male | 81 | 57 |
|  | Other | 0 | 0 |
|  | Did not specify | 1 | 1 |
| Race and/or ethnicity<br>(n = 148) | Asian | 7 | 0 |
|  | Black | 1 | 0 |
|  | Hispanic | 7 | 0 |
|  | Native American / American Indian | 0 | 0 |
|  | White | 127 | 86 |
|  | Other | 2 | 0 |
|  | Did not specify | 4 | 0 |

**Table 3.**
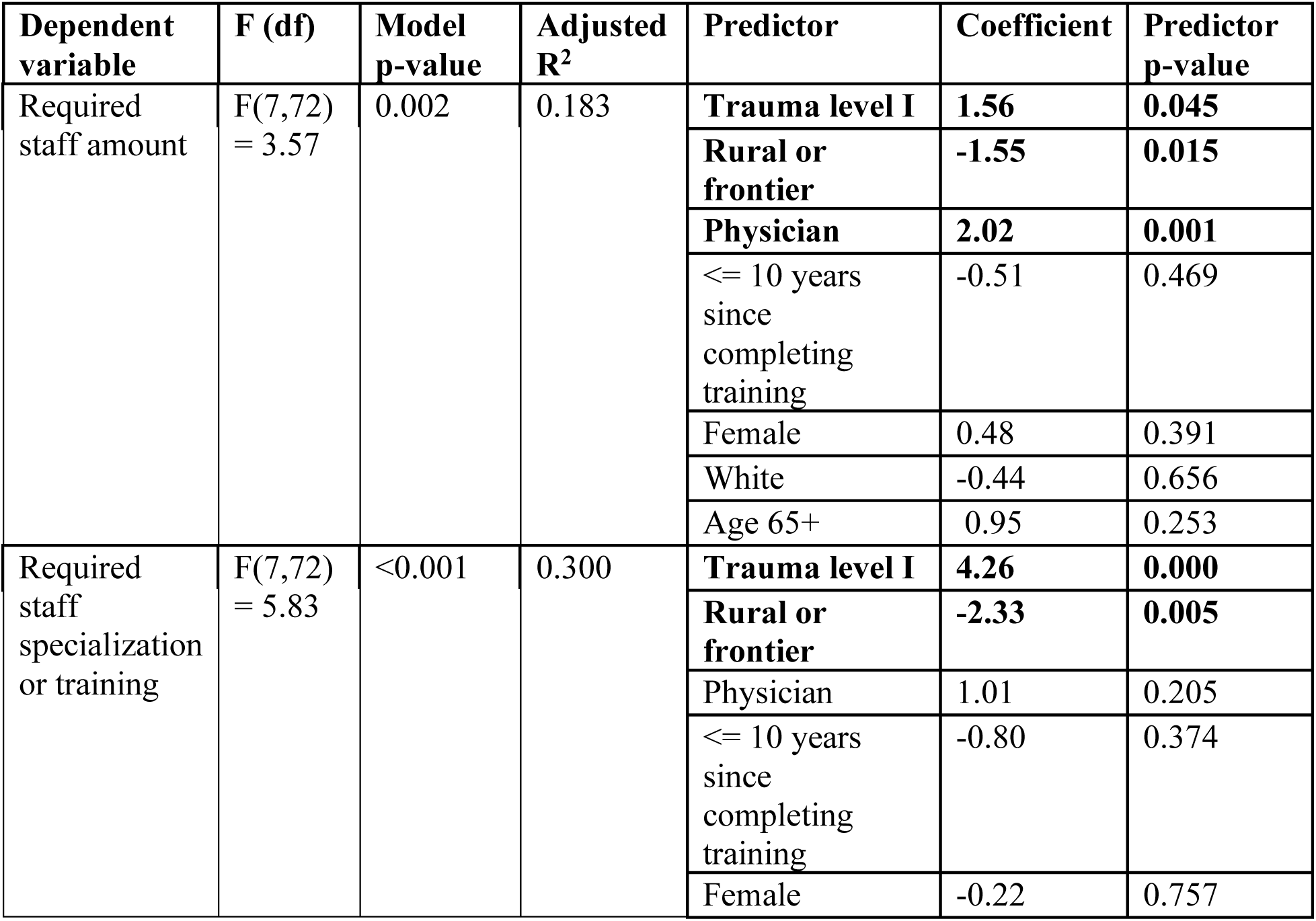
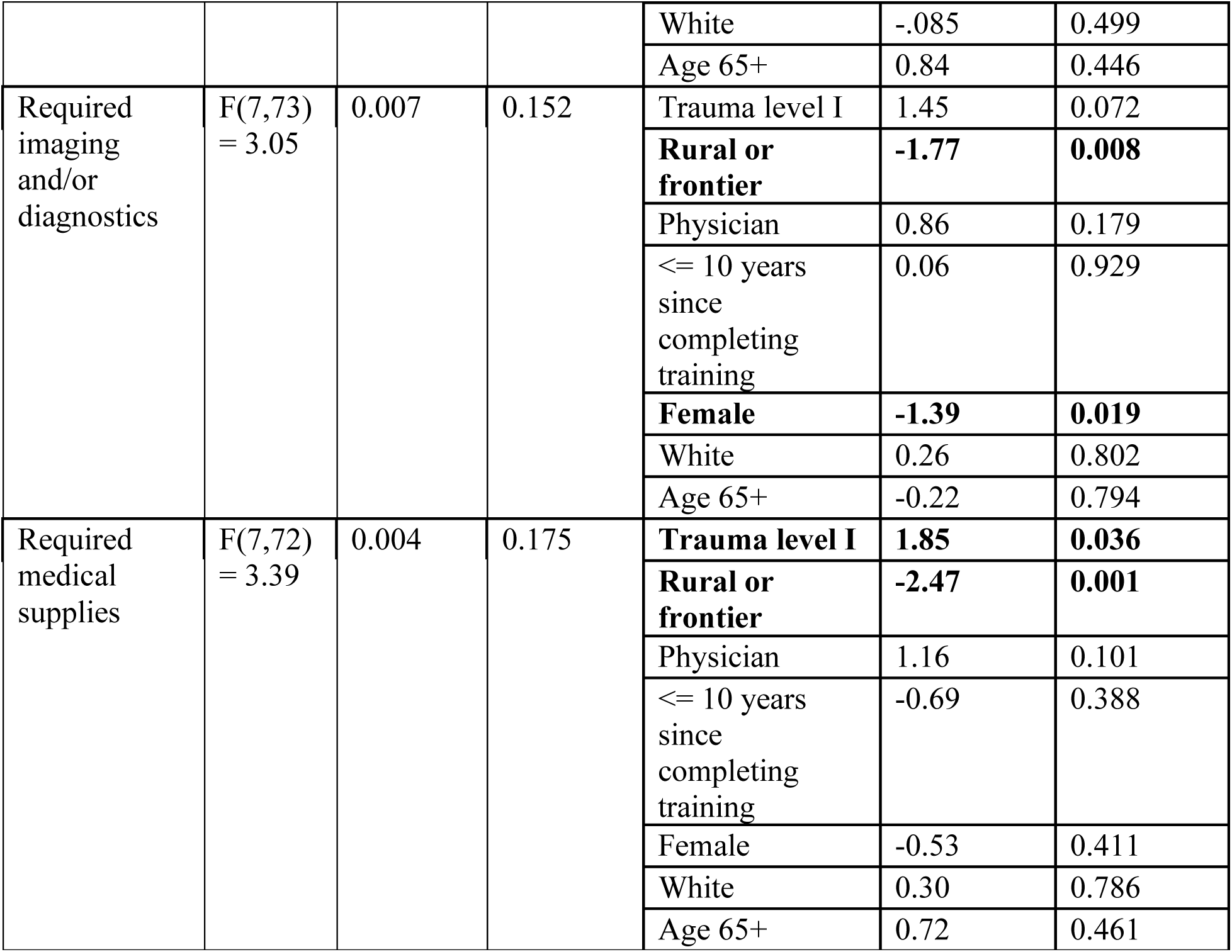
Multiple linear regression results for by resource type availability (significant results in bold).

| **Dependent variable** | **F (df)** | **Model p-value** | **Adjusted R<sup>2</sup>** | **Predictor** | **Coefficient** | **Predictor p-value** |
| --- | --- | --- | --- | --- | --- | --- |
| Required staff amount | F(7,72) = 3.57 | 0.002 | 0.183 | **Trauma level I** | **1.56** | **0.045** |
|  |  |  |  | **Rural or frontier** | **-1.55** | **0.015** |
|  |  |  |  | **Physician** | **2.02** | **0.001** |
|  |  |  |  | <= 10 years since completing training | -0.51 | 0.469 |
|  |  |  |  | Female | 0.48 | 0.391 |
|  |  |  |  | White | -0.44 | 0.656 |
|  |  |  |  | Age 65+ | 0.95 | 0.253 |
| Required staff specialization or training | F(7,72) = 5.83 | <0.001 | 0.300 | **Trauma level I** | **4.26** | **0.000** |
|  |  |  |  | **Rural or frontier** | **-2.33** | **0.005** |
|  |  |  |  | Physician | 1.01 | 0.205 |
|  |  |  |  | <= 10 years since completing training | -0.80 | 0.374 |
|  |  |  |  | Female | -0.22 | 0.757 |
|  |  |  |  | White | -.085 | 0.499 |
|  |  |  |  | Age 65+ | 0.84 | 0.446 |
| Required imaging and/or diagnostics | F(7,73) = 3.05 | 0.007 | 0.152 | Trauma level I | 1.45 | 0.072 |
|  |  |  |  | **Rural or frontier** | **-1.77** | **0.008** |
|  |  |  |  | Physician | 0.86 | 0.179 |
|  |  |  |  | <= 10 years since completing training | 0.06 | 0.929 |
|  |  |  |  | **Female** | **-1.39** | **0.019** |
|  |  |  |  | White | 0.26 | 0.802 |
|  |  |  |  | Age 65+ | -0.22 | 0.794 |
| Required medical supplies | F(7,72) = 3.39 | 0.004 | 0.175 | **Trauma level I** | **1.85** | **0.036** |
|  |  |  |  | **Rural or frontier** | **-2.47** | **0.001** |
|  |  |  |  | Physician | 1.16 | 0.101 |
|  |  |  |  | <= 10 years since completing training | -0.69 | 0.388 |
|  |  |  |  | Female | -0.53 | 0.411 |
|  |  |  |  | White | 0.30 | 0.786 |
|  |  |  |  | Age 65+ | 0.72 | 0.461 |

Little’s Test of Missingness revealed that data was not missing completely at random, and data was instead assumed to be missing at random. Missing data was handled with pairwise deletion during descriptive and regression analysis. Power analysis conducted in STATA showed a sample size need of 65 for multiple linear regression (alpha=0.05, power=0.8, number of predictor variables=7, target R-squared value=0.2). For binary logistic regression, data from previous workforce studies was used to estimate the proportion of participant characteristics (e.g., urban, physician) within the target population.^20,21^ Sample size needs were determined to be 105 to detect the desired odds ratio of 2.0. Mixed-method data from the interviews and surveys were compared using joint displays to identify similarities and differences in the results.^22^

## Results

Twelve participants completed semi-structured interviews (Table 1). 156 participants responded to the survey, 154 of whom were eligible, yielding a response rate of 4.8% (Table 2). All characteristics were self-reported. Participants working across multiple clinical sites were able to identify with multiple characteristics (e.g., trauma center level) in the interviews and survey. While completion of the demographic portions of the survey was relatively high (missing data rates of 0-14%), completion rates were considerably lower for the second set of instruments related to trauma clinical guidance needs (missing data rates of 47-57%). Analysis of missing data rates by participant characteristics did not reveal any consistent patterns.

### Current Usage of and Attitudes Towards Trauma Clinical Guidance

Clinicians reported using a variety of knowledge sources when facing clinical uncertainty, ranging from consults to decision-making support tools. Trauma clinical guidance was an important part of clinicians’ toolset for managing clinical uncertainty. Clinicians highlighted guidance’s ability to provide clarity and support for clinicians in stressful or unfamiliar scenarios. Clinicians emphasized that guidance helped standardize care, though guidance should never be a substitute for clinical judgement. When asked how often they used guidance in their clinical decision-making on a scale from never (0) to always (10), the median response was 7 (IQR=4-8, n=81). Participant characteristics were not significant predictors of guidance usage frequency.

### Barriers to Trauma Clinical Guidance Usage

The most cited barriers to guidance among participants include accessing guidance, guidance that is not up to date with the latest evidence, and resource insufficiency in practice settings. The median response to how often guidance was too hard to access at the time or place it was needed was 5 (0=never, 10=always, IQR=2-7, n=81). Interview participants shared their difficulties with finding guidance easily as it is dispersed across textbooks, national guidelines, printed posters or flyers, and other electronic resources. This fragmentation hinders use, with one physician stating, “If I have to work to find it, I may not use it” (P8). Clinicians reported a median score of 5 when asked if guidance is never (0) or always (10) up to date with the latest research findings (IQR=4-7, n=79). Despite acknowledging the work required to update guidance, a clinician still voiced frustration that “by the time the guideline actually gets updated and comes out, it’s old news” (P12).

Clinicians reported on a scale from 0 (never) to 10 (always) a median score of 5 when asked if their practice settings have the necessary resources to follow guidance recommendations (IQR=2-8, n=79). Multiple linear regression (F(7,73), p=0.002, adjusted R^2^=0.187) showed that practicing in a level I trauma center was a positive predictor for having the necessary resources to follow guidance (β=2.44, p=0.045). Practicing in a rural or frontier setting was a negative predictor for having necessary resources (β=-1.49, p=0.052).

Staffing limitations were more often the cause of resource constraints than physical resources. When asked if they never (0) or always (10) had the necessary resources when using guidance, clinicians reported a median of 5 for amount of staff (IQR=3-7, n=81), 5 for specialization of staff needed (e.g., surgeons) (IQR=0.5-8, n=80), 8 for imaging and diagnostics (IQR=6-9, n=81), and 8 for medical supplies such as blood products (IQR=4.5-9, n=80). Predictors for resource availability varied by resource type, though working at a level I trauma center or in a rural or frontier setting were common predictors of significance (Table 3).

Clinicians working outside urban and/or level 1 trauma centers frequently noted that trauma guidance resources, while thorough and detailing the best care currently available, are not relevant in their practice setting given their resource constraints. This lack of resources resulted in pivoting, either formally or informally, away from guidance recommendations. A rural clinician shared that “We only have two units of O+ and two units of O-…Our MTP is to transfer” (P2). Resource concerns extended beyond the rural setting; an urban, level III clinician (P8) described not having enough staff available to follow guidance.

Clinicians also reported other, less common but salient concerns about guidance barriers. Feeling too overwhelmed prevents guidance usage a median of 3 on a scale from 0 (never) to 10 (always) (IQR=1-5, n=81). Clinicians reported that formatting concerns prevent usage of guidance a median of 4 on the same scale (IQR=2-5, n=79), with one sharing that “if someone pulls up a guideline and it’s four pages long, they are turned off right away” (P1). Regression analysis did not demonstrate any participant characteristics to be significant predictors of guidance barriers outside of those already discussed with respect to resources constraints.

### Drivers of Guidance Usability

Guidance’s ability to be quickly understood in a time-pressured situation was the top driver of usability, with 47% of clinicians ranking it as the most important feature (n=70). Clinicians emphasized scarcity of time during trauma care, asserting that “Nothing is gonna stop for you to read a 20-page document. Nothing in the ED is gonna stop, particularly when you have these traumas…Time is of the essence…Time is against you” (P12). Many characteristics identified as vital to creating effective guidance were centered around usability in a time-pressured setting, including limiting the amount of text in a piece of guidance or employing flowchart design to streamline information.

Up-to-datedness was the second biggest driver of guidance usability, with 38% ranking it as the most important feature (n=66). Clinicians voiced frustration that guidance often lags the latest evidence, creating a need for clinicians to conduct their own evidence reviews. Other features of usability, including cost, customization to practice setting, and modality (e.g., mobile app vs EMR) were ranked lower relative to those top two priorities (Figure 3). Logistic regression exploring the effect of survey respondent characteristics on the likelihood of ranking any of these features as top two priorities did not yield any significant results.

**Figure 3.**
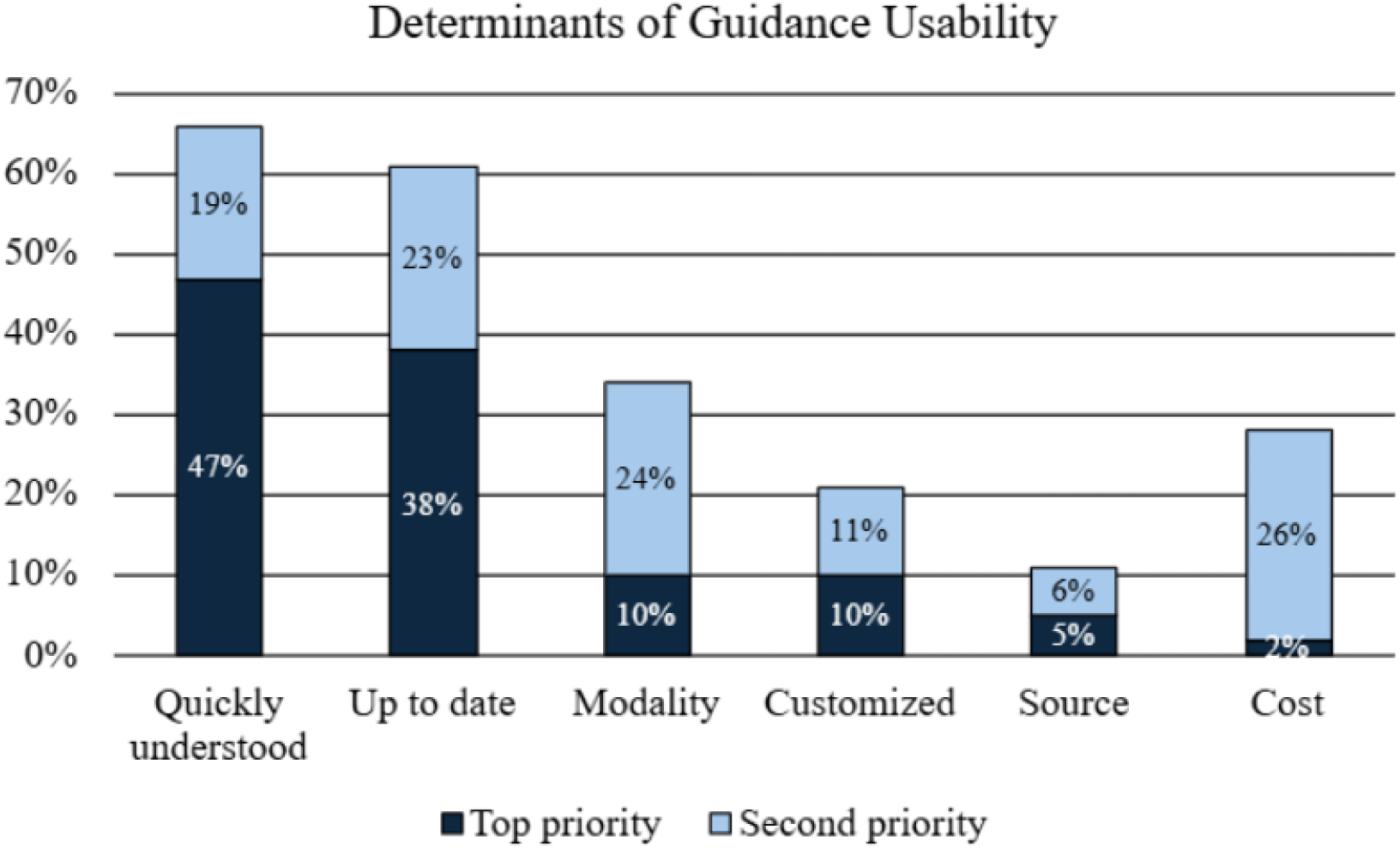
Determinants of guidance usability, frequency of ranking as top or second priority.

**Figure 4.**
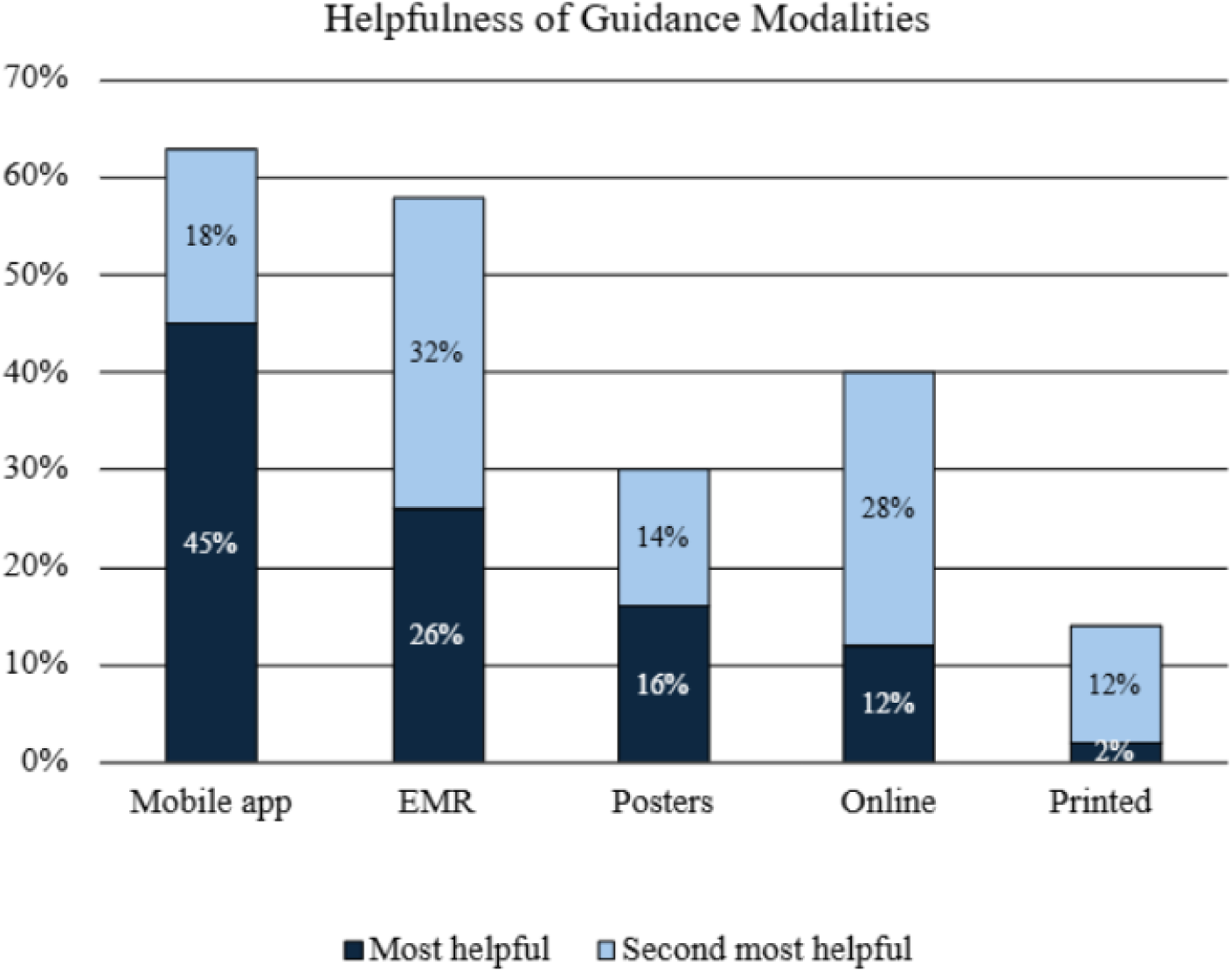
Helpfulness of guidance modalities, frequency of ranking as first or second most helpful

### Preferences for Guidance Modalities

Clinicians voiced a preference for guidance to be hosted primarily on electronic platforms, with mobile app and integration into the EMR identified as the most helpful guidance modalities. 45% of clinicians ranked mobile app as the most helpful modality (n=77). One clinician described mobile app as easy to access and that “95 percent of providers have their phones on them these days anyways” (P7). While clinicians emphasized ease of access as the main benefit of guidance delivery via mobile app, ease of translating guidance into patient care was the primary advantage for EMR-delivered guidance. EMR features such as clickable links to guidance embedded within order sets for blood units streamline the process of care while enabling clinicians to consult evidence during their decision-making process. 27% of clinicians ranked integration into the EMR as the most helpful modality (n=74). Some clinicians voiced preference for a multi-channel approach to guidance delivery, describing an ideal state where guidance could be found on mobile app, integrated into the EMR, and, for select pieces of high-frequency guidance (e.g., massive transfusion protocol), displayed on posters in a resuscitation bay. Regression did not identify any significant predictors of guidance modality preference.

### Preferences for Learning about Updates to Guidance

Clinicians described difficulty keeping up with frequent guidance changes, often receiving multiple updates via email each month that are difficult to internalize. Printed resources are inconsistently updated and timestamped, leaving clinicians unsure of their recency. Certain guidance modalities such as mobile app and EMR integration can reduce cognitive workload with automatic updates at the point of care. Otherwise, clinicians did not have a clear preference for learning about guidance updates (Figure 5).

**Figure 5.**
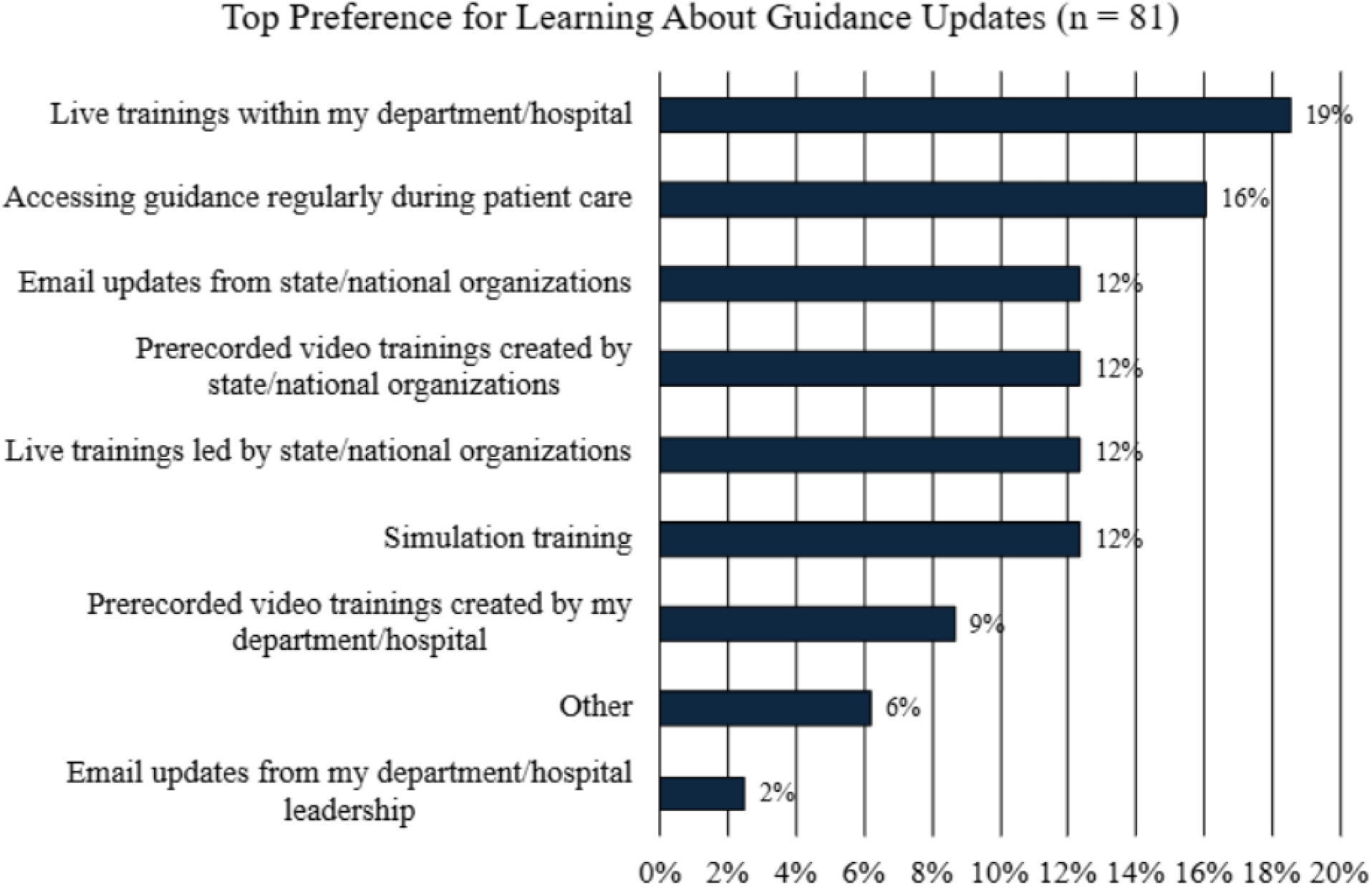
Top preferences for learning about guidance updates.

## Discussion

This work identifies gaps between existing trauma clinical guidance and the needs of emergency clinicians in their care of injured patients. Clinicians working in rural and frontier settings, as well as those working outside level 1 trauma centers, were significantly more likely to report that they lack sufficiently trained clinicians or resources to follow guidance recommendations. Providing recommendation options for lower-resource settings can eliminate barriers to implementation for clinicians working in those practice settings. Crafting guidance to be easily used at bedside and ensuring recommendations are up to date will improve guidance usability. Guidance should be delivered via preferred modalities such as mobile device applications, EMR, or a multi-modal approach to enable ease of access, application, and updates of guidance.

A main limitation in this study is the representativeness of the survey data of the overall emergency clinician workforce. The sample was collected through email outreach to members of EM professional associations and with a response rate of 4.8%, creating opportunity for response bias. The sample overrepresents certain groups (most notably APPs and white clinicians) and underrepresents others (rural) relative to recent workforce estimates, limiting generalizability of non-regression analyses (e.g., ranking the relative importance of guidance barriers or characteristics) to the broader emergency clinician population.^20,21^ Another limitation is the relatively small sample size. Although the number of responses generally met the sample size needs for the linear regression models, they did not consistently meet additional needs for the logistic regression models. The resulting under-powering of the survey may have resulted in type II error, particularly with respect to identifying predictors of specific guidance preferences.

Interview participants may not fully represent the diversity of emergency clinicians. Given that participants were known to the authorship group through professional networks, they may have been involved in prior efforts in guidance development. As a result, findings may reflect the perspective of more engaged stakeholders and may not capture the full range of experiences.

Potential next steps for this work include broadening distribution of the survey to ensure a more representative, robust sample, as well as user design testing with example guidance developed in line with the preferences of clinicians identified. These findings can inform development of future guidance by a range of key constituents in the trauma community, ranging from those writing original guidance recommendations at the global or national level to those adapting guidance for their local practice settings. Future scope could also be expanded to explore the role of artificial intelligence tools in guidance.

## Conclusion

The trauma community should continue to consider the variety of end users and clinical settings, including emergency clinicians, when developing trauma clinical guidance. Opportunities to better meet users’ needs, including developing guidance that is mobile-friendly and quickly understandable during patient care, should be a priority for authors of trauma clinical guidance.

## Supporting information

Supplemental Item 1

Supplemental Item 2

Supplemental Item 3

## Data Availability

All data produced in the present study are available upon reasonable request to the authors

**Supplemental Item 1** Consolidated criteria for reporting qualitative research (COREQ) Checklist.

**Supplemental Item 2** Checklist for Reporting Of Survey Studies (CROSS) Checklist.

**Supplemental Item 3** Survey instrument.

