## Supplemental Item 1 for "Assessing Emergency Clinicians’ Needs from Trauma Clinical Guidance: A Mixed-Methods Study"

| **No. Item** | **Guide description** | **Explanation** | **Reported on Page #** |
| --- | --- | --- | --- |
| **Domain 1: Research team and reflexivity** | | | |
| *Personal characteristics* | | | |
| 1. Interviewer / facilitator | Which authors conducted the interview or focus group? | Lindsay Fountain | 6 |
| 2. Credentials | What were the researchers’ credentials? | The project research team included diverse perspectives from the trauma and healthcare research communities, including three surgeons (JG, LL), a research physician (GZ), four emergency physicians (CK, JG, JW, NC), one emergency physician assistant (TM), one trauma nurse practitioner (AJ), one medical anthropologist (AN), and two medical students (KC, LF). | - |
| 3. Occupation | What was their occupation at the time of the study? | Medical student | 6 |
| 4. Gender | Was the researcher male or female? | Female | - |
| 5. Experience and training | What experience or training did the researcher have? | LF was trained in qualitative interviewing techniques by a medical anthropologist. | 6 |
| *Relationship with participants* | | | |
| 6. Relationship established | Was there a relationship established prior to study commencement? | No | 6 |
| 7. Participant knowledge of the interviewer | What did the participants know about the researcher? E.g. personal goals, reasons for doing the research | Participants were informed on the background and goals of the study before the interview | - |
| 8. Interviewer characteristics | What characteristics were reported about the interviewer/facilitator? e.g. Bias, assumptions, reasons and interests in the research topic | LF is a medical student with experience working as a scribe in two emergency departments | - |
| **Domain 2: Study design** | | | |
| *Theoretical framework* | | | |
| 9. Methodological orientation and theory | What methodological orientation was stated to underpin the study? e.g. grounded theory, discourse analysis, ethnography, phenomenology, content analysis | We conducted rapid qualitative content analysis of interview notes and audio recordings. | 6 |
| *Participant selection* | | | |
| 10. Sampling | How were participants selected? | Eligible participants included active EM clinicians working in the United States. Potential participants were initially identified using the coauthors’ professional networks. Purposeful criterion and snowball sampling further informed selection. | 4 |
| 11. Method of approach | How were participants approached? e.g. face-to-face, telephone, mail, email | Participants were contacted via email | 4-5 |
| 12. Sample size | How many participants were in the study? | 12 participants in total | 8 |
| 13. Non-participation | How many people refused to participate or dropped out? Reasons? | 22 participants who received our invitations did not respond or were not able to schedule an interview due to time constraints. | - |
| *Setting* | | | |
| 14. Setting of data collection | Where was the data collected? e.g. home, clinic, workplace | Data was collected via video conference | 5 |
| 15. Presence of non-participants | Was anyone else present besides the participants and researchers? | Interviews were conducted without anyone else present | - |
| 16. Description of sample | What are the important characteristics of the sample? e.g. demographic data, date | Described in Table 1 | 18 |
| *Data collection* | | | |
| 17. Interview guide | Were questions, prompts, guides provided by the authors? Was it pilot tested? | The interviews utilized a semi-structured format. Questions are shared in Supplemental Item 3. | - |
| 18. Repeat interviews | Were repeat interviews carried out? If yes, how many? | No | - |
| 19. Audio/visual recording | Did the research use audio or visual recording to collect the data? | All interviews were audio-recorded. | 6 |
| 20. Field notes | Were field notes made during and/or after the interview or focus group? | Yes, field notes were made during data collection and checked for accuracy by a secondary analyst during analysis. | 6 |
| 21. Duration | What was the duration of the interviews or focus group? | The average interview duration was 51 minutes, ranging from 31 to 63 minutes. | - |
| 22. Data saturation | Was data saturation discussed? | Information power was prioritized over data saturation. The sample size of twelve was sufficient given the high information power yielded by the study’s narrow aims, dense sample specificity, and strong dialogue. | 5 |
| 23. Transcripts returned | Were transcripts returned to participants for comment and/or correction? | No | - |
| **Domain 3: analysis and findings** | | | |
| *Data analysis* | | | |
| 24. Number of data coders | How many data coders coded the data? | Rapid qualitative methods were used, rather than traditional coding methodology. Two analysts reviewed the interview summary for each interview, then the primary analyst transferred findings into an Excel matrix for analysis. | 6 |
| 25. Description of coding tree | Did authors provide a description of the coding tree? | N/A, see above | - |
| 26. Derivation of themes | Were themes identified in advance or derived from the data? | Thematic analysis was not conducted. Domains regarding visual design were derived a posteriori. | 6 |
| 27. Software | What software, if applicable, was used to manage the data? | Microsoft Office Word and Excel (Version 2106, Build 14131.20332) | 6 |
| 28. Participant checking | Did participants provide feedback on the findings? | Participants did not provide feedback. | - |
| *Reporting* | | | |
| 29. Quotations presented | Were participant quotations presented to illustrate the themes/findings? Was each quotation identified? e.g. participant number | Yes, in the results | 7-11 |
| 30. Data and findings consistent | Was there consistency between the data presented and the findings? | Yes | 7-11 |
| 31. Clarity of major themes | Were major themes clearly presented in the findings? | Yes | 7-11 |
| 32. Clarity of minor themes | Were minor themes clearly presented in the findings? | No | - |
