## Supplemental Item 2 for "Assessing Emergency Clinicians’ Needs from Trauma Clinical Guidance: A Mixed-Methods Study"

| Section/topic | Item | Item description and notes | Reported on page # |
|---|---|---|---|
| **Title and abstract** | | | |
| Title and abstract | 1a | State the word “survey” along with a commonly used term in title or abstract to introduce the study’s design. | 1 |
| | 1b | Provide an informative summary in the abstract, covering background, objectives, methods, findings/results, interpretation/discussion, and conclusions. | 1 |
| **Introduction** | | | |
| Background | 2 | Provide a background about the rationale of study, what has been previously done, and why this survey is needed. | 3 |
| Purpose/aim | 3 | Identify specific purposes, aims, goals, or objectives of the study. | 3 |
| **Methods** | | | |
| Study design | 4 | Specify the study design in the methods section with a commonly used term (e.g., cross-sectional or longitudinal). | 6 |
| | 5a | Describe the questionnaire (e.g., number of sections, number of questions, number and names of instruments used).<br>*The survey included a consent form, 16 items related to participant demographics, and 30 items related to trauma clinical guidance needs.* | See notes |
| Data collection methods | 5b | Describe all questionnaire instruments that were used in the survey to measure particular concepts. Report target population, reported validity and reliability information, scoring/classification procedure, and reference links (if any). | 4, 6, Supplemental Item 3 |
| | 5c | Provide information on pretesting of the questionnaire, if performed (in the article or in an online supplement). Report the method of pretesting, number of times questionnaire was pre-tested, number and demographics of participants used for pretesting, and the level of similarity of demographics between pre-testing participants and sample population.<br>*The survey was pre-tested in two rounds, with two medical students providing feedback in each round. The students involved in pretesting differed from survey participants in their more limited medical knowledge base and younger age.* | See notes |
| | 5d | Questionnaire if possible, should be fully provided (in the article, or as appendices or as an online supplement). | Supplemental Item 3 |

| | | | |
|---|---|---|---|
| Sample characteristics | 6a | Describe the study population (i.e., background, locations, eligibility criteria for participant inclusion in survey, exclusion criteria). | 4 |
| | 6b | Describe the sampling techniques used (e.g., single stage or multistage sampling, simple random sampling, stratified sampling, cluster sampling, convenience sampling). Specify the locations of sample participants whenever clustered sampling was applied. | 4 |
| | 6c | Provide information on sample size, along with details of sample size calculation. | 7-8 |
| | 6d | Describe how representative the sample is of the study population (or target population if possible), particularly for population-based surveys. | 12 |
| Survey administration | 7a | Provide information on modes of questionnaire administration, including the type and number of contacts, the location where the survey was conducted (e.g., outpatient room or by use of online tools, such as SurveyMonkey). | 4,6 |
| | 7b | Provide information of survey's time frame, such as periods of recruitment, exposure, and follow-up days.<br>*Surveys were shared once via email with no follow up recruitment. The survey remained open for responses for 60 days.* | See notes |
| | 7c | Provide information on the entry process:<br>–>For non-web-based surveys, provide approaches to minimize human error in data entry.<br>–>For web-based surveys, provide approaches to prevent "multiple participation" of participants.<br>*There were not specific measures in place to prevent multiple participation of participants, though line-by-line review of survey data did not show data trends suggestive of significant incidents of multiple participation (e.g., repeated identical entries).* | See notes |
| Study preparation | 8 | Describe any preparation process before conducting the survey (e.g., interviewers' training process, advertising the survey).<br>*The interviewer (LF) completed a live online course in semi-structured interview techniques ahead of conducting interviews. The interview guide, summary template, and matrix were reviewed by a methodological expert (AN) prior to usage.* | See notes |

| | | | |
|---|---|---|---|
| Ethical considerations | 9a | Provide information on ethical approval for the survey if obtained, including informed consent, institutional review board [IRB] approval, Helsinki declaration, and good clinical practice [GCP] declaration (as appropriate). | 4 |
| | 9b | Provide information about survey anonymity and confidentiality and describe what mechanisms were used to protect unauthorized access. | 4,6 |
| Statistical analysis | 10a | Describe statistical methods and analytical approach. Report the statistical software that was used for data analysis. | 6-7 |
| | 10b | Report any modification of variables used in the analysis, along with reference (if available). | 7, Table 3 |
| | 10c | Report details about how missing data was handled. Include rate of missing items, missing data mechanism (i.e., missing completely at random [MCAR], missing at random [MAR] or missing not at random [MNAR]) and methods used to deal with missing data (e.g., multiple imputation). | 7 |
| | 10d | State how non-response error was addressed. | 7 |
| | 10e | For longitudinal surveys, state how loss to follow-up was addressed. | NA |
| | 10f | Indicate whether any methods such as weighting of items or propensity scores have been used to adjust for non-representativeness of the sample. | NA |
| | 10g | Describe any sensitivity analysis conducted. | NA |
| **Results** | | | |
| Respondent characteristics | 11a | Report numbers of individuals at each stage of the study. Consider using a flow diagram, if possible.<br>*See response to 11c for the number of participants at each stage of the study. The number of participants who viewed the survey but did not start it was not captured.* | See notes |
| | 11b | Provide reasons for non-participation at each stage, if possible. | NA |
| | 11c | Report response rate, present the definition of response rate or the formula used to calculate response rate.<br>*The overall response rate was 4.8%, with 154 eligible respondents from a pool of 1,546 advanced practice providers and 1,630 physicians* | 7; see notes for additional detail |

| | | | |
|---|---|---|---|
| | 11d | Provide information to define how unique visitors are determined. Report number of unique visitors along with relevant proportions (e.g., view proportion, participation proportion, completion proportion).<br><br>*The number of unique visitors was not able to be determined.* | See notes |
| Descriptive results | 12 | Provide characteristics of study participants, as well as information on potential confounders and assessed outcomes. | Table 2 |
| Main findings | 13a | Give unadjusted estimates and, if applicable, confounder-adjusted estimates along with 95% confidence intervals and p-values. | 7-11 |
| | 13b | For multivariable analysis, provide information on the model building process, model fit statistics, and model assumptions (as appropriate). | 7-11 |
| | 13c | Provide details about any sensitivity analysis performed. If there are considerable amount of missing data, report sensitivity analyses comparing the results of complete cases with that of the imputed dataset (if possible). | NA |
| **Discussion** | | | |
| Limitations | 14 | Discuss the limitations of the study, considering sources of potential biases and imprecisions, such as non-representativeness of sample, study design, important uncontrolled confounders. | 12 |
| Interpretations | 15 | Give a cautious overall interpretation of results, based on potential biases and imprecisions and suggest areas for future research. | 12-13 |
| Generalizability | 16 | Discuss the external validity of the results. | 12-13 |
| **Other sections** | | | |
| Role of funding source | 17 | State whether any funding organization has had any roles in the survey's design, implementation, and analysis.<br><br>*No funding organization participated in the survey process.* | See notes |
| Conflict of interest | 18 | Declare any potential conflict of interest.<br><br>*No conflicts of interest to declare.* | See notes |
| Acknowledgements | 19 | Provide names of organizations/persons that are acknowledged along with their contribution to the research. | NA |
