## Supplemental Item 3 for "Assessing Emergency Clinicians’ Needs from Trauma Clinical Guidance: A Mixed-Methods Study"

At what type of hospital location do you primarily work? (check all that apply)

- ☐ Urban
- ☐ Suburban
- ☐ Rural
- ☐ Frontier
- ☐ Other

If other, please specify.

__________________________________

What is the trauma center level designation at your primary hospital? (check all that apply)

- ☐ Level I
- ☐ Level II
- ☐ Level III
- ☐ Level IV
- ☐ Level V
- ☐ Other/unknown

If other/unknown, please explain.

__________________________________

What is the estimated transport time from your hospital to the nearest level I trauma center?

- ○ Less than 30 minutes
- ○ 30-59 minutes
- ○ 1-2 hours
- ○ 3+ hours

In which state is your primary hospital located?

- ◯ AK
- ◯ AL
- ◯ AR
- ◯ AZ
- ◯ CA
- ◯ CO
- ◯ CT
- ◯ DC
- ◯ DE
- ◯ FL
- ◯ GA
- ◯ HI
- ◯ IA
- ◯ ID
- ◯ IL
- ◯ IN
- ◯ KS
- ◯ KY
- ◯ LA
- ◯ MA
- ◯ MD
- ◯ ME
- ◯ MI
- ◯ MN
- ◯ MO
- ◯ MS
- ◯ MT
- ◯ NC
- ◯ ND
- ◯ NE
- ◯ NH
- ◯ NJ
- ◯ NM
- ◯ NV
- ◯ NY
- ◯ OH
- ◯ OK
- ◯ OR
- ◯ PA
- ◯ PR
- ◯ RI
- ◯ SC
- ◯ SD
- ◯ TN
- ◯ TX
- ◯ UT
- ◯ VA
- ◯ VT
- ◯ WA
- ◯ WI
- ◯ WV
- ◯ WY

---

Which of the following clinician types do you most identify with?

- ◯ Emergency Medicine Attending Physician
- ◯ Emergency Medicine Nurse Practicioner
- ◯ Emergency Medicine PA
- ◯ Emergency Medicine Resident Physician
- ◯ Other

---

If other, please specify.

__________________________________

| | |
|---|---|
| How many years ago did you complete your clinical training? | ◯ 0-5 years<br>◯ 6-10 years<br>◯ 11-20 years<br>◯ 21+ years<br>◯ Still in training |
| Select your credentials (check all that apply) | ☐ MD<br>☐ DO<br>☐ PhD<br>☐ DNP<br>☐ MSN<br>☐ PA-C<br>☐ RN<br>☐ MA<br>☐ MS<br>☐ MPH<br>☐ Other (specify) |
| If other, please specify. | __________________________________ |
| What best describes your gender identity? | ◯ Woman<br>◯ Man<br>◯ Transwoman<br>◯ Transman<br>◯ Non-binary / non-conforming<br>◯ Prefer not to respond<br>◯ Prefer to self-describe (specify) |
| Please specify. | __________________________________ |
| Which race and/or ethnicity do you identify with? (select all that apply) | ☐ Native American/American Indian<br>☐ Hispanic/Latino<br>☐ Asian<br>☐ Black or African descent<br>☐ White<br>☐ Prefer not to respond<br>☐ Other (specify) |
| If other, please specify. | __________________________________ |
| What is your age? | ◯ Less than 25 years<br>◯ 25-34 years<br>◯ 35-44 years<br>◯ 45-54 years<br>◯ 55-64 years<br>◯ 65+ years |

### Trauma Clinical Guidance Needs

Please complete the survey below.

Thank you!

#### Please indicate how often the following impact your decision to use trauma clinical guidance.

| | 0 = never | 1 | 2 | 3 | 4 | 5 | 6 | 7 | 8 | 9 | 10 (always) |
|---|---|---|---|---|---|---|---|---|---|---|---|
| I _____ use trauma clinical guidance in my clinical decision making | ○ | ○ | ○ | ○ | ○ | ○ | ○ | ○ | ○ | ○ | ○ |
| I am _______ too overloaded to use trauma clinical guidance. | ○ | ○ | ○ | ○ | ○ | ○ | ○ | ○ | ○ | ○ | ○ |
| Trauma clinical guidance is _______ too hard to access at the time/place that I need them (e.g. consider paywalls, technological barriers, difficulty locating the guidance). | ○ | ○ | ○ | ○ | ○ | ○ | ○ | ○ | ○ | ○ | ○ |
| Trauma clinical guidance formatting ________ makes the guidance too cumbersome to use. | ○ | ○ | ○ | ○ | ○ | ○ | ○ | ○ | ○ | ○ | ○ |
| My practice setting _______ has the necessary resources (e.g., materials, staff) to follow the clinical guidance recommendation(s). | ○ | ○ | ○ | ○ | ○ | ○ | ○ | ○ | ○ | ○ | ○ |
| Trauma clinical guidance recommendations are _______ consistent with the most up-to-date research findings. | ○ | ○ | ○ | ○ | ○ | ○ | ○ | ○ | ○ | ○ | ○ |

Optional: Include any additional comments to help explain your responses. __________________________________

#### How important are the following features in deciding trauma clinical guidance usability? Rank the following options.

| | 1 (most important) | 2 | 3 | 4 | 5 | 6 (least important) |
|---|---|---|---|---|---|---|
| Current with most up-to-date research findings | ○ | ○ | ○ | ○ | ○ | ○ |
| Cost effective (e.g. free or affordable) | ○ | ○ | ○ | ○ | ○ | ○ |

| | | | | | | |
|---|---|---|---|---|---|---|
| Available in my preferred modality (e.g., mobile app, built into electronic medical record) | ○ | ○ | ○ | ○ | ○ | ○ |
| Quickly understandable in a time-pressured situation | ○ | ○ | ○ | ○ | ○ | ○ |
| Customized to different resource settings (e.g., different levels of trauma centers) | ○ | ○ | ○ | ○ | ○ | ○ |
| Source and/or authorship | ○ | ○ | ○ | ○ | ○ | ○ |

Optional: Include why these features are important to you and any additional comments. ______

**Which methods for accessing trauma clinical guidance are most helpful? Rank the following options.**

| | 1 (most helpful) | 2 | 3 | 4 | 5 (least helpful) |
|---|---|---|---|---|---|
| Standalone mobile app | ○ | ○ | ○ | ○ | ○ |
| Integrated into electronic medical record | ○ | ○ | ○ | ○ | ○ |
| Centralized online resource | ○ | ○ | ○ | ○ | ○ |
| Displayed on posters and/or signs inside the unit | ○ | ○ | ○ | ○ | ○ |
| Collected in printed resource (e.g., binder) inside the unit | ○ | ○ | ○ | ○ | ○ |

Optional: Include why these methods are helpful for you and any additional comments. ______

| | 0 (rarely) | 1 | 2 | 3 | 4 | 5 | 6 | 7 | 8 | 9 | 10 (always) |
|---|---|---|---|---|---|---|---|---|---|---|---|
| When using trauma clinical guidance, we _______ have the amount of staff needed . | ○ | ○ | ○ | ○ | ○ | ○ | ○ | ○ | ○ | ○ | ○ |
| When using trauma clinical guidance, we ________ have the training/specialization of staff needed (e.g., trauma surgeons) | ○ | ○ | ○ | ○ | ○ | ○ | ○ | ○ | ○ | ○ | ○ |
| When using trauma clinical guidance, we _______ have the imaging and/or diagnostics needed. | ○ | ○ | ○ | ○ | ○ | ○ | ○ | ○ | ○ | ○ | ○ |
| When using trauma clinical guidance, we _____ have the medical supplies needed (e.g., chest tube trays, blood products) | ○ | ○ | ○ | ○ | ○ | ○ | ○ | ○ | ○ | ○ | ○ |

Optional: Include what resource limitations you face in your practice setting and any additional comments. ______

| | |
|---|---|
| I prefer to learn about updates to trauma clinical guidance through | ◯ Live trainings within my department/hospital<br>◯ Prerecorded video trainings created by my department/hospital<br>◯ Simulation training<br>◯ Email updates from my department/hospital leadership<br>◯ Live trainings led by state/national organizations<br>◯ Prerecorded video trainings created by state/national organizations<br>◯ Email updates from state/national organizations<br>◯ Accessing the guidance regularly during patient care (e.g., a electronic resource that automatically updates)<br>◯ Other [write-in] |
| If other, please specify. | ______________________ |
| What would you change about trauma clinical guidance to make it more accessible and relevant in your context? | ______________________ |
| Please share any other thoughts you have about improving trauma clinical guidance for emergency medicine clinicians. | ______________________ |
| Thank you for completing this survey. We value your perspective and will use this information to make improvements to trauma clinical guidance. If you would like to be contacted in the future regarding potential research panels or interviews, please leave your email address below. | ______________________ |
